# Intrinsic Signal Amplification by Type-III CRISPR-Cas Systems Provides a Sequence-Specific Viral Diagnostic

**DOI:** 10.1101/2020.10.14.20212670

**Authors:** Andrew Santiago-Frangos, Laina N. Hall, Anna Nemudraia, Artem Nemudryi, Pushya Krishna, Tanner Wiegand, Royce A. Wilkinson, Deann T. Snyder, Jodi F. Hedges, Mark A. Jutila, Matthew P. Taylor, Blake Wiedenheft

## Abstract

To combat viral pandemics, there is an urgent need for inexpensive new technologies that enable fast, reliable, and scalable detection of viruses. Here we repurposed the type III CRISPR-Cas system for sensitive and sequence specific detection of SARS-CoV-2 in an assay that can be performed in one hour or less. RNA recognition by type III systems triggers Cas10-mediated polymerase activity, which simultaneously generates pyrophosphates, protons and cyclic oligonucleotides. We show that amplified products of the Cas10-polymerase are detectable using colorimetric or fluorometric readouts.

---

Frequent tests and quick results are critical for stopping the spread of SARS-CoV-2 and ending the current COVID-19 pandemic (Larremore et al., 2020; Paltiel et al., 2020). RT-qPCR (reverse transcriptase-quantitative polymerase chain reaction) has been the gold standard for viral diagnostics, but this method is slow and requires sophisticated equipment that is expensive to purchase and operate. Thus, there is an urgent need for inexpensive new technologies that enable fast, reliable, and scalable detection of viruses.

Recently, loop-mediated isothermal amplification (LAMP) (Notomi et al., 2000) has been developed as a sensitive (1-100 copies/μL) point-of-care diagnostic (Dao Thi et al., 2020; Zhang et al., 2020). However, LAMP is prone to generating false positives unless a second sequence-specific technique is used to check the amplified DNA (Dao Thi et al., 2020; Rolando et al., 2020). The type V (Cas12-based) and type IV (Cas13-based) CRISPR systems have recently been coupled to LAMP or RPA (recombinase polymerase amplification) for sensitive and reliable detection of viral nucleic acids (Chen et al., 2018; Gootenberg et al., 2018). Following isothermal amplification, the RNA-guided Cas12 or Cas13 proteins bind to the amplified target and trigger a non-sequence specific nuclease activity that cleaves a fluorophore and quencher labelled DNA or RNA (Chen et al., 2018; Gootenberg et al., 2018). Cleavage of the tether results in an increase in fluorescence that can be detected in 45 minutes (**Supplemental Table 1**).

While Cas12 and Cas13 detection methods have been optimized over several iterations to be compatible with isothermal amplification of viral RNA, we wondered if the intrinsic signal amplification mechanisms that are unique to and conserved in the type III CRISPR-Cas systems could eliminate or minimize the need for upstream nucleic acid amplification prior to CRISPR-based detection (**Fig. 1**). Here, we show that the Csm complex from *Thermus thermophilus* (TtCsm) can be programed to specifically recognize the SARS-CoV-2 genome. SARS-CoV-2, but not SARS-CoV-1, activates the Cas10 polymerase, which generates ∼1000 cyclic nucleotides (e.g., cA_4_) after binding an RNA target (Jia et al., 2019; Kazlauskiene et al., 2017; Niewoehner et al., 2017; Rouillon et al., 2018). Like all polymerases, nucleotide polymerization by Cas10, also generates protons (H^+^) and pyrophosphates (PPi). We demonstrate that each of these three products can be used to detect SARS-CoV-2 RNA using either colorimetric, fluorometric, or both methods simultaneously. The assay can be performed in 10 to 45 minutes depending on the detection method and the concentration of the RNA. While direct detection of RNA using the type III Csm complex from *T. thermophilus* is 100-fold more sensitive than direct detection by Cas13 (Cas12 only binds dsDNA), an initial nucleic acid amplification step is needed for detection of viral RNA in patient samples and further optimization is required to improve the compatibility of Csm-based detection with LAMP.

**Figure 1.**
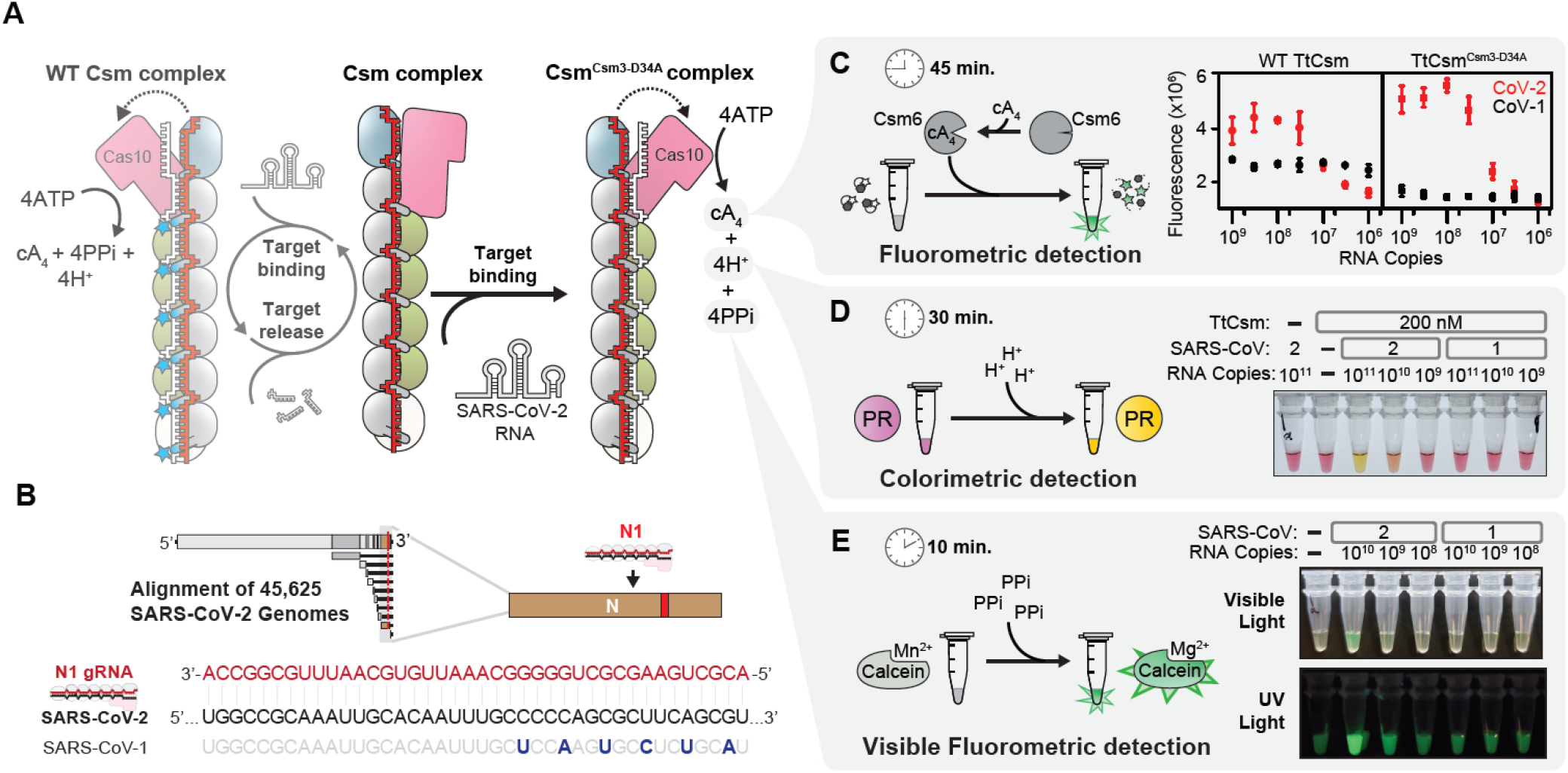
Detection of SARS-CoV-2 using the type III CRISPR-Cas system. (**A**) Schematic of the type III Csm complex from *Thermus thermophilus* (TtCsm), and enzymatic activities triggered by binding to a complementary RNA target. TtCsm complex consists of a single crRNA (red) and an unequal stoichiometry of five different proteins (Cas10_1_, pink: Csm4_1_, blue: Csm3_6_, grey: Csm2_4_, green: Csm5_1_, white). RNA binding activates the Cas10-polymerase, Cas10 DNase, and Csm3 RNase (left, transparent). Target RNA cleavage by Csm3 subunits (blue stars) causes target dissociation and inactivates Cas10. A mutation in the Csm3 subunit (TtCsm^Csm-D34A^) renders the complex RNase-dead (right). (**B**) Schematic of SARS-CoV-2 genome. The CRISPR RNA (crRNA) targets a sequence on the 3’ end of genomic and subgenomic RNAs, that is conserved in 45,625 SARS-CoV-2 genomes (i.e., 99.99% of all available genomes). Mismatches between the crRNA-guide and SARS-CoV-1 are highlighted (navy blue). (**C**) Fluorometric detection of *in vitro* transcribed SARS-CoV-2 (red squares) and SARS-CoV-1 N-gene (black squares). Cyclic tetra-adenylate (cA_4_) activates TtCsm6. Activated TtCsm6 cleaves an RNA tether, which links a fluorophore (star) to a quencher (gray hexagon). TtCsm^Csm3-D34A^ (right graph) exhibits a LoD 3-fold lower than wildtype TtCsm (left graph) and retains specificity for SARS-CoV-2 RNA. The mean of three technical replicates are shown, error bars represent ± 1 standard deviation. (**D**) Colorimetric detection of SARS-CoV-2 RNA by TtCsm^Csm3-D34A^ complex using a pH sensitive dye (i.e., Phenol Red). Reactions were incubated for 30 minutes at 60°C. Technical replicates are shown (**Supplemental Fig. 3**). (**E**) Visible fluorometric detection of SARS-CoV-2 RNA by TtCsm^Csm3-D34A^ complex using Calcein. Reactions were incubated for up to an hour at 60°C. Technical replicates and kinetics are shown (**Supplemental Fig. 4**).

## Results and Discussion

### Sequence-specific activation of Cas10 polymerase yields three detectable products

Sequence-specific recognition of RNA by type III CRISPR systems initiates a signaling cascade (**Fig. 1A**) (Kazlauskiene et al., 2017; Niewoehner et al., 2017; Rouillon et al., 2018). RNA binding by the TtCsm complex triggers a conformational change that activates the Palm domain of the Cas10 subunit, which amplifies the RNA binding signal by converting ATP into approximately 1,000 cyclic oligoadenylates (e.g. cA_4_) (Jia et al., 2019; Kazlauskiene et al., 2017; Niewoehner et al., 2017; Rouillon et al., 2018). We hypothesized that the intrinsic signal amplification unique to type III CRISPR systems would boost the sensitivity of direct RNA detection, while maintaining specificity. To test this hypothesis, we expressed and purified the type III-A CRISPR RNA (crRNA)-guided surveillance complex from Thermus thermophilus (TtCsm) with a guide complementary to the N-gene of SARS-CoV-2 (**Fig. 1B and Supplemental Fig. 1**).

The Csm3 subunits, which form the “backbone” of the Csm complex, are nucleases that cleave bound target RNA in 6-nt increments (Liu et al., 2017; Samai et al., 2015; Tamulaitis et al., 2014). The cleaved RNA fragments dissociate and the Csm complex returns to the “inactive” state (i.e., no Cas10-polymerase activity) (**Fig. 1A**) (Nasef et al., 2019; Rouillon et al., 2018). Thus, in the context of an immune response, the RNase activity of Csm3 moderates Cas10 polymerase activity to limit excess nuclease activation (Athukoralage et al., 2020; Nasef et al., 2019; Rouillon et al., 2018), that may otherwise kill the cell. However, we reasoned that a Csm3 mutation that prevents target RNA degradation, would have two related benefits as a diagnostic. First, an RNase-dead Csm complex is expected to stay bound to target RNA longer, which would sustain the Cas10 polymerase activity. Second, Csm3-mediated cleavage of the target RNA (e.g., SARS-CoV-2 RNA) would reduce the target RNA concentration over time and thus limit the sensitivity of the assay. Therefore, we mutated residues in the Csm3 subunit responsible for target RNA cleavage (D34A) (Liu et al., 2017; Tamulaitis et al., 2014), and purified the RNase-dead complex (TtCsmCsm3-D34A) (**Supplemental Fig. 1**). To measure the limit of detection (LoD), we added the mutant or wildtype Csm complex to a reaction containing the TtCsm6 nuclease, a fluorescent reporter (i.e., FAM-RNA-Iowa Black FQ), and a titration of RNA corresponding to the N-gene of either SARS-CoV-2 or SARS-CoV-1 (**Fig. 1C and Supplemental Fig. 2**). Using fluorometric detection, both the mutant and the wildtype Csm complex could detect the SARS-CoV-2 RNA at concentrations above 10^8^ copies per reaction, and neither complex cross-reacted with the SARS-CoV-1 RNA at the highest concentrations tested. The RNase-dead TtCsm complex was roughly 3-fold more sensitive than wildtype, with an LoD of 10^7^ copies (3×10^8^ copies/mL).

In addition to fluorometric detection, we also developed a colorimetric RNA detection method that utilizes a pH change that occurs during nucleotide polymerization (**Fig. 1D and Supplemental Fig. 3**). Specific recognition of SARS-CoV-2 by RNase-dead TtCsm complex, activates Cas10. Cas10 polymerizes ATP (Jia et al., 2019; Kazlauskiene et al., 2017; Niewoehner et al., 2017; Rouillon et al., 2018), releasing one proton per incorporated nucleotide. Cas10-generated protons acidify the solution and change the color of a pH indicator (i.e. Phenol Red) from fuchsia through orange (10^10^ RNA copies), to yellow (10^11^ RNA copies).

Further, we developed a visible fluorometric RNA detection method that relies on the sequestration of metallic ions by pyrophosphate. The metal indicator Calcein is initially quenched by bound Mn^2+^ ions (Tomita et al., 2008). Specific binding of SARS-CoV-2 by RNase-dead TtCsm complex activates Cas10, which generates one pyrophosphate per ATP polymerized. Pyrophosphate forms an insoluble precipitate with Mn2+, which unquenches Calcein. Free Calcein is then bound by excess Mg^2+^, forming a highly fluorescent complex that can be seen by eye or with a UV lamp (**Fig. 1E**). 10^10^ copies of SARS-CoV-2 RNA were detected in 10 minutes at 60°C (**Supplemental Fig. 4**).

### Coupled RT-LAMP-T7-Csm detects SARS-CoV-2 infections

The LoD of type III CRISPR-based fluorometric detection (**Fig. 1C**) is roughly 100-10,000 fold better than direct RNA or DNA detection by Cas12, Cas13 or Cascade (**Supplemental Table 1**) (Chen et al., 2018; Gootenberg et al., 2018; Yoshimi et al., 2020). However, it is currently not sensitive enough to directly detect SARS-CoV-2 in all patients capable of spreading the infection, which requires detecting 10^6^ RNA copies/mL (La Scola et al., 2020; Larremore et al., 2020; Paltiel et al., 2020; Wölfel et al., 2020). To further decrease the LoD of a type III CRISPR-based diagnostic to 10^6^ copies/mL or lower, we incorporated an upstream nucleic acid amplification technique (**Fig. 2A**). First, SARS-CoV-2 genomic RNA is reverse transcribed into DNA (RT), which is then amplified by LAMP. We designed LAMP primers that incorporate a T7 promoter into DNA products, which are then transcribed by thermostable T7 RNA polymerase, into RNA targets that are detected by the TtCsm complex. To validate this method, we tested RNA extracted from 24 nasopharyngeal swab samples taken from patients that had previously been tested using RT-qPCR (**Fig. 2B**). Of the 24 samples tested, 16 were positive for SARS-CoV-2, seven were negative and one was inconclusive (the Ct for N2 marker was <40, but N1 was not detected). For these samples, RT-LAMP-T7-Csm has a specificity (negative predictive agreement) of 100%, as well as a positive predictive agreement of 100% for nasopharyngeal swab samples with more than 10^6^ copies/mL, which is a minimal infectious concentration in cell culture (La Scola et al., 2020; Wölfel et al., 2020). Patient Sample 1 was inconclusive for both RT-qPCR and RT-LAMP-T7-Csm. RT-LAMP-T7-Csm is specific for SARS-CoV-2 and is sufficiently sensitive to identify viral titers of 10^6^ copies/mL in a 1-hour reaction (**Supplemental Fig. 5**).

**Figure 2.**
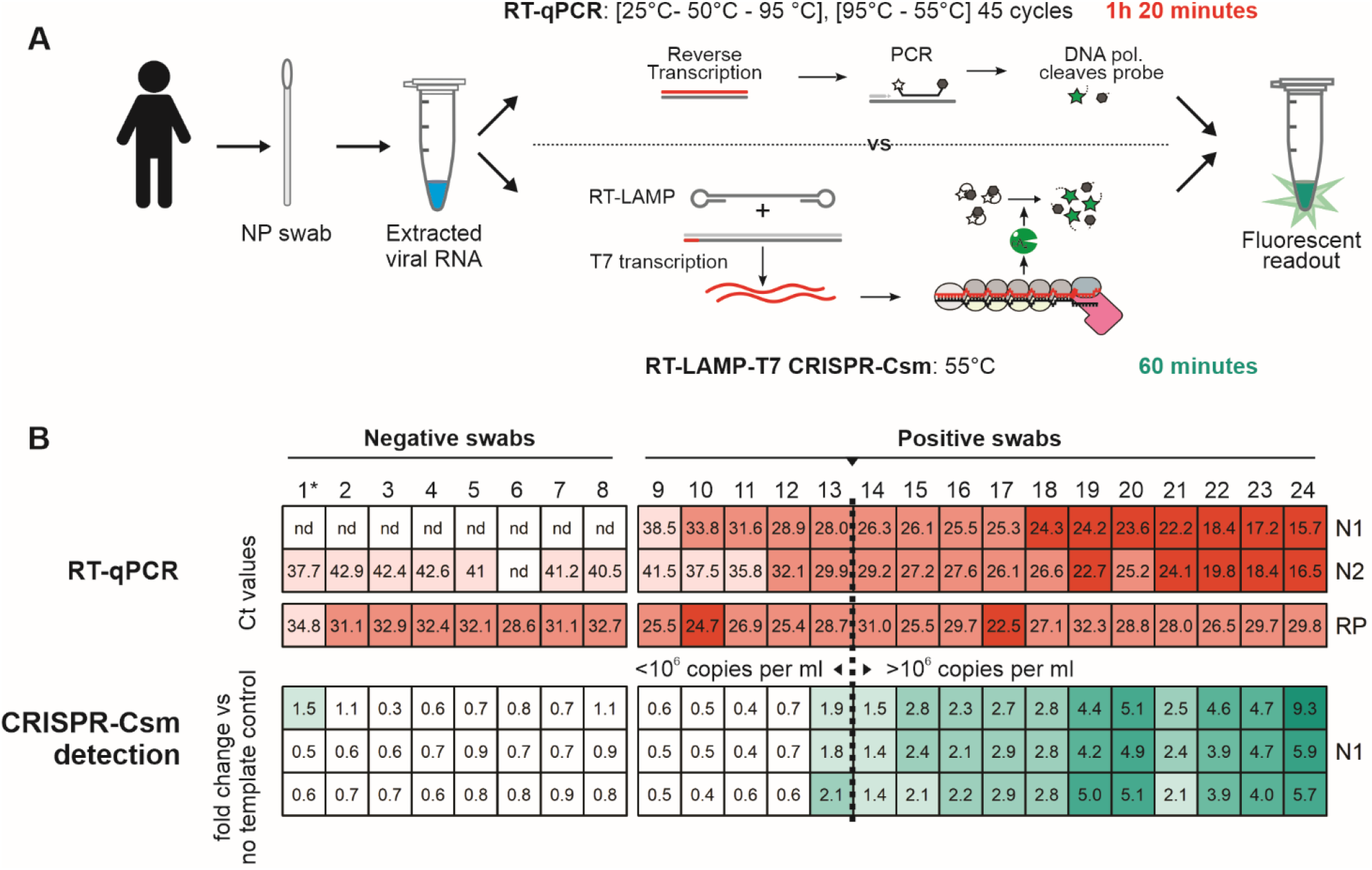
CRISPR-Csm-based detection of SARS-CoV-2 in clinical nasopharyngeal swab samples. (**A**) Schematic comparing RT-qPCR to RT-LAMP-CRISPR-Csm based detection. *In RT-qPCR reaction*, reverse transcribed template is amplified in PCR reaction that cycles between different temperatures. As Taq DNA polymerase (DNA pol.) synthesizes nascent DNA strand it degrades the probe annealed to amplified template and releases the fluorophore (green star) from the quencher (black hexagon). *In CRISPR-Csm detection*, the viral RNA is reverse transcribed, and resulting DNA is amplified in an RT-LAMP reaction to produce transcription templates for T7 RNA polymerase, in one pot. An aliquot of the RT-LAMP-T7 reaction (40 min) is then mixed with CRISPR-Csm reaction (20 min). (**B**) Nasopharyngeal swabs from 24 individuals were tested with RT-qPCR and RT-LAMP-T7 CRISPR-Csm. *Top panel* shows threshold cycle (Ct) values for RT-qPCR. Swabs with Ct values below 40 for both N1 and N2 CDC diagnostic primers are considered positive for SARS-CoV-2 RNA (nd – signal is not detected over during 45 cycles). Vertical dashed line highlights the Ct value corresponding to a concentration of 10^6^ SARS-CoV-2 RNA copies/mL (**Supplemental Fig. 4**). *Bottom panel* shows CRISPR-Csm based detection in the same swab samples targeting nucleocapsid (N) gene, as shown in (A). Data is shown as fold change in fluorescence compared to a no-template control reaction. Technical repeats are shown in triplicate. * - swab sample test result was inconclusive both for CDC assay and CRISPR-Csm detection.

Collectively, this work demonstrates that sequence specific detection of viral RNA by the type III CRISPR-Cas complex triggers the synthesis of cA_4_, pyrophosphate, and protons, each of which are detectable within 10 to 45 minutes, using colorimetric or fluorometric methods (**Fig. 1**). Additional work is needed to improve the sensitivity of type III CRISPR-based direct RNA detection without upstream amplification (e.g., LAMP, RPA, etc.). Current improvements are focused on increasing the homogeneity of the TtCsm complex (**Supplemental Fig. 1**) and screening for crRNAs that result in high specificity and high signal. Guides that meet these performance criteria will increase sensitivity and may enable use of type III CRISPR systems without prior amplification.

## Materials and Methods

### Nucleic acid preparation

Previously published LAMP primers (Eurofins) were designed to amplify the SARS-CoV-2 N-gene (Broughton et al., 2020). Target SARS-CoV-2 and SARS-CoV-1 RNAs were *in vitro* transcribed with MEGAscript T7 (Thermo Fisher Scientific) from PCR products generated from pairs of synthesized overlapping DNA oligos (**Supplemental Table 2**) (Eurofins), as per the manufacturer’s instructions. Transcribed RNAs were purified by denaturing PAGE. Fluorescent reporter RNA (/56-FAM/rCrUrCrUrCrU/3IABkFQ/) was purified by RNase-free HPLC (IDT).

### Plasmids

Expression vectors for *Thermus thermophilus* type III-A *csm1-csm5* genes, pCDF-5xT7-TtCsm were purchased from Addgene (plasmid # 128572) (Liu et al., 2019). pCDF-5xT7-TtCsm was used as a template for site-directed mutagenesis to mutate the Csm3 residue D33 to alanine (D33A) to inactivate Csm3-mediated cleavage of target RNA (pCDF-5xT7-TtCsm^Csm3-D34A^) (Liu et al., 2017). The CRISPR array in pACYC-TtCas6-4xcrRNA4.5 (Addgene plasmid # 127764) (Liu et al., 2019) was replaced with a synthetic CRISPR array (GeneArt) containing five repeats and four identical spacers, designed to target the N-gene of SARS-CoV2 (i.e., pACYC-TtCas6-4xgCoV2N1). The nuclease TtCsm6 was expressed from pC0075 TtCsm6 His6-TwinStrep-SUMO-BsaI (Addgene plasmid # 115270) (Gootenberg et al., 2018).

### Protein purifications

Expression and purification of the TtCsm complex was performed as previously described with minor modifications (Liu et al., 2019). Briefly, pACYC-TtCas6-4xgCoV2N1 was co-transformed with either pCDF-5xT7-TtCsm or pCDF-5xT7-TtCsm^Csm3-D34A^, into *Escherichia coli* BL21(DE3) cells and grown in LB Broth (Lennox) (Thermo Fisher Scientific), at 37°C to an OD_600_ of 0.5. Cultures were then incubated on ice for 1 hour, and then induced with 0.5 mM IPTG (isopropyl-β-D-thiogalactoside), for expression overnight at 16°C. Cells were lysed via sonication, in Lysis buffer (25 mM HEPES pH 7.5, 150 mM KCl, 10 mM imidazole, 1 mM TCEP, 0.01 % Triton X-100, 5 % glycerol, 1 mM PMSF) and Halt Protease Inhibitor Cocktail (Thermo Fisher Scientific) and lysate was clarified by centrifugation at 15,000 rpm, for 30 mins at 4°C. The lysate was then heat-treated at 55°C for 45 minutes and further clarified by centrifugation at 15,000 rpm, for 30 mins at 4°C. His-tagged Csm1 and TtCsm complex were bound to Ni-NTA resin (Qiagen) and washed with Wash buffer (50 mM HEPES pH 7.5, 150 mM KCl, 1 mM TCEP, 5 % glycerol, 20 mM imidazole, 2 mM ATP and 10 mM MgCl_2_). Protein was eluted in Lysis buffer supplemented with 300 mM imidazole. Eluted protein was concentrated (Corning Spin-X concentrators) at 4°C before further purification over a HiLoad Superdex 200 26/600 size-exclusion column (Cytiva) in 25 mM HEPES pH 7.5, 150 mM NaCl, 5% glycerol, 1 mM TCEP. Fractions containing the TtCsm complex were pooled, aliquoted and stored at −80°C.

Expression and purification of TtCsm6 was performed as previously described with minor modifications (Gootenberg et al., 2018). pTtCsm6 was transformed into *Escherichia coli* BL21(DE3) cells and grown in LB Broth (Lennox) (Thermo Fisher Scientific), at 37°C to an OD600 of 0.5. Cultures were then incubated on ice for 1 hour, and then induced with 0.5 mM IPTG (isopropyl-β-D-thiogalactoside), for expression overnight at 16°C. Cells were lysed via sonication, in TtCsm6 Lysis buffer (20 mM Tris-HCl pH 8, 500 mM NaCl, 1 mM DTT) and Halt Protease Inhibitor Cocktail (Thermo Scientific) and lysate was clarified by centrifugation at 15,000 rpm, for 30 mins at 4°C. The lysate was heat-treated at 55°C for 45 minutes, and clarified by centrifugation at 15,000 rpm, for 30 mins at 4°C. TwinStrep-tagged TtCsm6 was bound to StrepTrap HP resin (Cytiva) and washed in TtCsm6 Lysis buffer. The protein was eluted with TtCsm6 Lysis buffer supplemented with 2.5 mM desthiobiotin and concentrated (Corning Spin-X concentrators) at 4°C. Affinity tags were removed from TtCsm6 using SUMO protease (100 μL of 2.5 mg/ml protease per 20 mg of TtCsm6 substrate) in SUMO digest buffer (30 mM Tris-HCl pH 8, 500 mM NaCl 1 mM DTT, 0.15% Igepal) at 4°C overnight. The digest was run over the StrepTrap HP resin, and the flow-through was concentrated using Corning Spin-X concentrators at 4°C. Finally, TtCsm6 was purified using a HiLoad Superdex 200 26/600 size-exclusion column (Cytiva) in 20 mM Tris-HCl pH 7.5, 1 mM DTT, 400 mM Monopotassium glutamate, 5 % Glycerol. Fractions containing TtCsm6 were pooled, aliquoted and stored at −80°C.

### Type III CRISPR-based RNA detection

#### Fluorescent CRISPR-Csm based detection

RNA was extracted from nasopharyngeal swabs derived from patients that tested negative for SARS-CoV-2 as determined by RT-qPCR. This RNA was used as is or spiked with *in vitro* transcribed SARS-CoV-2 or SARS-CoV-1 RNA. These RNA samples were mixed with 250 μM ATP, 500 nM fluorescent reporter RNA, 500 nM TtCsm complex, and 2500 nM of TtCsm6 in reaction buffer (20 mM Tris-HCl pH 7.9, 200 mM Monopotassium glutamate, 10 mM Ammonium sulphate, 5 mM Magnesium sulphate and 1 mM TCEP (tris(2-carboxyethyl)phosphine)) in a 30 μL reaction. Reactions were incubated at 60°C (CRISPR-Csm alone), and fluorescence was measured over time in an ABI 7500 Fast Real-Time PCR System (Applied Biosystems), using the manufacturers default filter settings for FAM dye. Fluorescence measurements at an incubation time of 45 minutes are reported.

#### Colorimetric CRISPR-Csm based detection

TtCsm^Csm3-D34A^ stocks were buffer exchanged into a low buffering capacity buffer (0.5 mM Tris-HCl pH 8.8, 50 mM Potassium chloride, 10 mM Ammonium sulphate, 8 mM Magnesium sulphate) using Microspin G25 columns (Cytiva) as per the manufacturer’s instructions. TE buffer (10 mM Tris-HCl pH 7.5, 1 mM EDTA) or *in vitro* transcribed SARS-CoV-2 or SARS-CoV-1 RNA were incubated with 200 nM TtCsm^Csm3-D34A^ in 1x WarmStart Colorimetric LAMP Master Mix (NEB), supplemented with an additional 1 mM ATP, in a 25 μL reaction. The volume of buffer-exchanged TtCsm used contributed approximately 40 μM Tris-HCl pH 8.8 buffer to the final reaction. Reactions were assembled on ice and imaged on an LED tracing pad with a Galaxy S9 phone (Samsung). Then reactions were incubated at 60°C for 30 minutes, rapidly cooled, and imaged again.

#### Visible fluorometric CRISPR-Csm based detection

TE buffer or *in vitro* transcribed SARS-CoV-2 or SARS-CoV-1 RNA were incubated with 500 nM TtCsm^Csm3-D34A^ in reaction buffer (20 mM Tris-HCl pH 8.8, 100 mM Potassium chloride, 10 mM Ammonium sulphate, 6 mM Magnesium sulphate, 0.5 mM Manganese chloride, 1 mM TCEP, 1 mM ATP and 25 μM Calcein), in a 30 μL reaction. Reactions were incubated at 60°C, and fluorescence was measured over time in an ABI 7500 Fast Real-Time PCR System (Applied Biosystems), using the manufacturers default filter settings for FAM dye. After incubating at 60°C for 50 minutes, the same reactions were then imaged under visible light, and under UV light (365 nm) with a Galaxy S9 phone (Samsung).

### RT-LAMP-T7 CRISPR-Csm

Isothermal amplification of nucleic acids in swab samples was performed by combining RT-LAMP (NEB) with *in vitro* transcription in a single reaction using Hi-T7 RNA polymerase (NEB). In brief, 25 μl reactions contained 8 units (U) of WarmStart Bst 2.0 (NEB), and 7.5 U of WarmStart RTx Reverse Transriptase (NEB), 1.4 mM dNTPs, LAMP primers, 125 U of Hi-T7 (NEB), 0.5 mM rNTPs, 25 U of Murine RNase Inhibitor (NEB) in isothermal amplification buffer (20 mM Tris-HCl pH 7.8, 8 mM Magnesium sulfate, 10 mM Ammonium sulfate, 50 mM potassium chloride, 0.1% Tween-20). LAMP primers designed to amplify the SARS-CoV-2 N-gene (Broughton et al., 2020), were added at a final concentration of 0.2 μM F3 and B3, 0.4 μM LoopF and LoopB, 1.6 μM FIP and BIP, and 0.8 μM of T7-FIP. The T7-FIP primer consists of a T7 promoter fused to the 3’ half of the FIP primer, and allows for the generation of T7 transcription templates during LAMP (**Supplemental Table 1**). Reactions were performed using 1 μl of input RNA at 55°C for 40 minutes. 20 μl of RT-LAMP-T7 reactions were mixed with 30 µl of a modified CRISPR-Csm fluorescent detection reaction: 250 μM ATP, 500 nM fluorescent reporter RNA, 100 nM TtCsm complex, and 100 nM of TtCsm6 in reaction buffer (20 mM Tris-HCl pH 7.8, 2 mM Magnesium sulfate, 10 mM Ammonium sulfate, 50 mM potassium chloride, 0.1% Tween-20). Reactions were incubated at 55°C for 20 min and fluorescence kinetics was monitored with ABI 7500 Fast Real-Time PCR System.

### Human clinical sample collection and preparation

Nasopharyngeal swabs from patients that either tested negative or positive for SARS-CoV-2 were collected in viral transport media. RNA was extracted from all patient samples using QIAamp Viral RNA Mini Kit (Qiagen).

### RT-qPCR

RT-qPCR was performed using two primers pairs (N1 and N2) and probes from the 2019-nCoV CDC EUA Kit (IDT#10006606). SARS-CoV-2 in RNA-extracted, nasopharyngeal patient samples was detected and quantified using one-step RT-qPCR in ABI 7500 Fast Real-Time PCR System according to CDC guidelines and protocols (https://www.fda.gov/media/134922/download). In brief, 20 µL reactions included 8.5 µL of Nuclease-free Water, 1.5 µL of Primer and Probe mix (IDT, 10006713), 5 µL of TaqPath 1-Step RT-qPCR Master Mix (ThermoFisher, A15299) and 5 µL of the template. Nuclease-free water was used as negative template control (NTC). Amplification was performed as follows: 25°C for 2 min, 50°C for 15 min, 95 °C for 2 min followed by 45 cycles of 95 °C for 3 s and 55 °C for 30 s. To quantify viral genome copy numbers in the samples, standard curves for N1 and N2 were generated using a dilution series of a SARS-CoV-2 synthetic RNA fragment (RTGM 10169, National Institute of Standards and Technology) spanning N gene with concentrations ranging from 10 to 10^6^ copies per μL. Three technical replicates were performed at each dilution. The NTC showed no amplification throughout the 45 cycles of qPCR.

### Bioinformatic analyses

45,652 SARS-CoV-2 genomes defined as complete (>29,000 bp) were downloaded from GISAID (https://www.gisaid.org/) (Elbe and Buckland-Merrett, 2017), on June 23, 2020. Sequences were aligned by GISAID using MAFFT’s auto-alignment settings (Katoh and Standley, 2013). Sequences from animals (e.g. pangolin, bat) and those containing ambiguous sequencing data (e.g. N’s) were removed. The alignments were used to design the N1 TtCsm crRNA which specifically targets a sequence amplified by the RT-LAMP primers (described above). 78 (0.17%) SARS-CoV-2 genomes contain a single mismatch (77 genomes) or four mismatches (1 genome) to the first 18 nucleotides of N1, but it is unlikely that a single mismatch in this region is sufficient to prevent Csm-mediated viral detection. Thus, the N1 guide is expected to target 99.99% of published SARS-CoV-2 sequences. The generation and viewing of viral genome alignments was performed using publicly available software, including SnapGene Viewer and RStudio V1.2.5001 (Methods).

### Statistical analyses

All experiments were performed in triplicate and error is reported as ± 1 standard deviation.

## Data Availability

The data shown in the manuscript are available upon request from the corresponding author. Expression plasmids are available upon request. SARS-CoV-2 genomes were accessed on the GISAID database website (https://www.gisaid.org/).

## Acknowledgements

We are grateful to members of Bozeman Health that provided deidentified patient samples. Specifically, Christopher Nero, Douglas Smoot, Winfield Wallace, Cayley Faurot-Daniels, Verena Lawrence and Melissa Blauvelt. We are also grateful to Michelle Flenniken, Katie Daughenbaugh, Diane Bimczok and other members of the COVID task force at MSU for assistance establishing the COVID testing center. A.S.-F. is a postdoctoral fellow of the Life Science Research Foundation, which is supported by the Simons Foundation. A.S.-F. is supported by the Postdoctoral Enrichment Program Award from the Burroughs Wellcome Fund. Research in the Wiedenheft lab and the biosafety level 3 facility is supported by the office of the Vice President for Research at Montana State University, and a sponsored research agreement from VIRIS Detection Systems Inc.

## Author Contributions

B.W. conceived the experimental plan. A.S.-F., and R.A.W. conducted fluorometric and colorimetric Csm-based RNA detection. A.S.-F conducted visual fluorometric Csm-based RNA detection. L.N.H., P.K. and R.A.W., purified proteins. A. Nemudraia, A. Nemudryi performed RT-qPCR. A.S.-F., A. Nemudraia and A. Nemudryi performed RT-LAMP-T7-Csm-based RNA detection. T.W. performed bioinformatic analyses and designed crRNA guides. D.T.S., and J.F.H. performed RNA extraction of patient nasopharyngeal swab samples under the guidance of M.A.J. and M.P.T.. A.S.-F., T. W., A. Nemudraia, A. Nemudryi, and B.W. wrote the manuscript.

## References

Athukoralage JS, Graham S, Rouillon C, Grüschow S, Czekster CM, White MF. 2020. The dynamic interplay of host and viral enzymes in type iii crispr-mediated cyclic nucleotide signalling. Elife 9:1–16. doi:10.7554/eLife.55852

Broughton JP, Deng X, Yu G, Fasching CL, Servellita V, Singh J, Miao X, Streithorst JA, Granados A, Sotomayor-Gonzalez A, Zorn K, Gopez A, Hsu E, Gu W, Miller S, Pan C, Guevara H, Wadford DA, Chen JS, Chiu CY. 2020. CRISPR–Cas12-based detection of SARS-CoV-2. Nat Biotechnol. doi:10.1038/s41587-020-0513-4

Chen JS, Ma E, Harrington LB, Da Costa M, Tian X, Palefsky JM, Doudna JA. 2018. CRISPR-Cas12a target binding unleashes indiscriminate single-stranded DNase activity. Science (80-) 360:436–439. doi:10.1126/science.aar6245

Dao Thi VL, Herbst K, Boerner K, Meurer M, Kremer LP, Kirrmaier D, Freistaedter A, Papagiannidis D, Galmozzi C, Stanifer ML, Boulant S, Klein S, Chlanda P, Khalid D, Miranda IB, Schnitzler P, Kräusslich H-G, Knop M, Anders S. 2020. A colorimetric RT-LAMP assay and LAMP-sequencing for detecting SARS-CoV-2 RNA in clinical samples. Sci Transl Med 7075:1–20. doi:10.1126/scitranslmed.abc7075

Elbe S, Buckland-Merrett G. 2017. Data, disease and diplomacy: GISAID’s innovative contribution to global health. Glob Challenges 1:33–46. doi:10.1002/gch2.1018

Gootenberg JS, Abudayyeh OO, Kellner MJ, Joung J, Collins JJ, Zhang F. 2018. Multiplexed and portable nucleic acid detection platform with Cas13, Cas12a and Csm6. Science (80-) 360:439–444. doi:10.1126/science.aaq0179

Jia N, Jones R, Sukenick G, Patel DJ. 2019. Second Messenger cA4 Formation within the Composite Csm1 Palm Pocket of Type III-A CRISPR-Cas Csm Complex and Its Release Path. Mol Cell 75:933-943.e6. doi:10.1016/j.molcel.2019.06.013

Katoh K, Standley DM. 2013. MAFFT Multiple Sequence Alignment Software Version 7: Improvements in Performance and Usability. Mol Biol Evol 30:772–780. doi:10.1093/molbev/mst010

Kazlauskiene M, Kostiuk G, Venclovas Č, Tamulaitis G, Siksnys V. 2017. A cyclic oligonucleotide signaling pathway in type III CRISPR-Cas systems. Science (80-) 357:605–609. doi:10.1126/science.aao0100

La Scola B, Le Bideau M, Andreani J, Hoang VT, Grimaldier C, Colson P, Gautret P, Raoult D. 2020. Viral RNA load as determined by cell culture as a management tool for discharge of SARS-CoV-2 patients from infectious disease wards. Eur J Clin Microbiol Infect Dis 39:1059–1061. doi:10.1007/s10096-020-03913-9

Larremore DB, Wilder B, Lester E, Shehata S, Burke JM, Hay JA, Tambe M, Mina MJ, Parker R. 2020. Test sensitivity is secondary to frequency and turnaround time for COVID-19 surveillance. medRxiv 2020.06.22.20136309. doi:10.1101/2020.06.22.20136309

Liu TY, Iavarone AT, Doudna JA. 2017. RNA and DNA targeting by a reconstituted Thermus thermophiles Type III-A CRISPR-Cas system. PLoS One 12:1–20. doi:10.1371/journal.pone.0170552

Liu TY, Liu JJ, Aditham AJ, Nogales E, Doudna JA. 2019. Target preference of Type III-A CRISPR-Cas complexes at the transcription bubble. Nat Commun 10. doi:10.1038/s41467-019-10780-2

Nasef M, Muffly MC, Beckman AB, Rowe SJ, Walker FC, Hatoum-Aslan A, Dunkle JA. 2019. Regulation of cyclic oligoadenylate synthesis by the Staphylococcus epidermidis Cas10-Csm complex. Rna 25:948–962. doi:10.1261/rna.070417.119

Niewoehner O, Garcia-Doval C, Rostøl JT, Berk C, Schwede F, Bigler L, Hall J, Marraffini LA, Jinek M. 2017. Type III CRISPR–Cas systems produce cyclic oligoadenylate second messengers. Nature 548:543–548. doi:10.1038/nature23467

Notomi T, Okayama H, Masubuchi H, Yonekawa T, Watanabe K, Amino N, Hase T. 2000. Loop-mediated isothermal amplification of DNA. Nucleic Acids Res 28:E63. doi:10.1093/nar/28.12.e63

Paltiel AD, Zheng A, Walensky RP. 2020. Assessment of SARS-CoV-2 Screening Strategies to Permit the Safe Reopening of College Campuses in the United States. JAMA Netw open 3:e2016818. doi:10.1001/jamanetworkopen.2020.16818

Rolando JC, Jue E, Barlow JT, Ismagilov RF. 2020. Real-time kinetics and high-resolution melt curves in single-molecule digital LAMP to differentiate and study specific and non-specific amplification. Nucleic Acids Res 48:e42. doi:10.1093/nar/gkaa099

Rouillon C, Athukoralage JS, Graham S, Grüschow S, White MF. 2018. Control of cyclic oligoadenylate synthesis in a type III CRISPR system. Elife 7:1–22. doi:10.7554/eLife.36734

Samai P, Pyenson N, Jiang W, Goldberg GW, Hatoum-Aslan A, Marraffini LA. 2015. Co-transcriptional DNA and RNA cleavage during type III CRISPR-cas immunity. Cell 161:1164–1174. doi:10.1016/j.cell.2015.04.027

Tamulaitis G, Kazlauskiene M, Manakova E, Venclovas Č, Nwokeoji AO, Dickman MJ, Horvath P, Siksnys V. 2014. Programmable RNA Shredding by the Type III-A CRISPR-Cas System of Streptococcus thermophilus. Mol Cell 56:506–517. doi:10.1016/j.molcel.2014.09.027

Tomita N, Mori Y, Kanda H, Notomi T. 2008. Loop-mediated isothermal amplification (LAMP) of gene sequences and simple visual detection of products. Nat Protoc 3:877–882. doi:10.1038/nprot.2008.57

Wölfel R, Corman VM, Guggemos W, Seilmaier M, Zange S, Müller MA, Niemeyer D, Jones TC, Vollmar P, Rothe C, Hoelscher M, Bleicker T, Brünink S, Schneider J, Ehmann R, Zwirglmaier K, Drosten C, Wendtner C. 2020. Virological assessment of hospitalized patients with COVID-2019. Nature 581:465–469. doi:10.1038/s41586-020-2196-x

Yoshimi K, Takeshita K, Yamayoshi S, Shibumura S, Yamauchi Y, Yamamoto M, Yotsuyanagi H, Kawaoka Y, Mashimo T. 2020. Rapid and accurate detection of novel coronavirus SARS-CoV-2 using CRISPR-Cas3. medRxiv 2020.06.02.20119875. doi:10.1101/2020.06.02.20119875

Zhang Y, Ren G, Buss J, Barry AJ, Patton GC, Tanner NA. 2020. Enhancing colorimetric loop-mediated isothermal amplification speed and sensitivity with guanidine chloride. Biotechniques 69:1–8. doi:10.2144/btn-2020-0078

